# Prevalence and Relationship of Rest Tremor and Action Tremor in Parkinson’s Disease

**DOI:** 10.1101/2020.11.09.20227850

**Authors:** Deepak K. Gupta, Massimo Marano, Cole Zweber, James T. Boyd, Sheng-Han Kuo

## Abstract

**Background:** Despite the significance of tremor in Parkinson’s disease (PD) diagnosis, classification, and patient’s quality of life, there is a relative lack of data on prevalence and relationship of different tremor types in PD.

**Methods:** The presence of rest tremor (RT) and action tremor (AT; defined as combination of both postural and kinetic tremor) was determined and RT severity was defined using the Movement Disorders Society Unified Parkinson’s Disease Rating Scale (MDS-UPDRS) at baseline in the Progression Marker Initiative (PPMI, n=423), the Fox Investigation for New Discovery of Biomarkers (BioFIND, n=118) and the Parkinson’s Disease Biomarkers Program (PDBP, n=873) cohorts.

**Results:** Across baseline data of all three cohorts, RT prevalence (58.2%) was higher than AT prevalence (39.0%). Patients with RT had significantly higher (Chi-square test, p<0.05) prevalence of AT compared to patients without RT in the PPMI (40.0% versus 30.1%), BioFIND (48.0% versus 40.0%) and PDBP (49.9% versus 21.0%) cohorts. Furthermore, patients with AT had significantly (Student t-test, p<0.05) higher RT severity that those without AT in PPMI (5.7 ± 5.4 versus 3.9 ± 3.3), BioFIND, 6.4 ± 6.3 versus 3.8 ± 4.4) and PDBP (6.4 ± 6.6 versus 3.7 ± 4.4) cohorts.

**Discussion:** The RT is the most frequent tremor type and present in more than half of the PD patients. However, AT is also present in nearly one-third of the PD patients. Our results also indicate that RT and AT may have cross-interactions in PD.

## Introduction

Tremor has been defined as a rhythmic and oscillatory involuntary movement with varied phenomenology, which can be classified based on several parameters, such as body parts, frequency and activation state[1]. The latest consensus classification by the Movement Disorders Society (MDS) classifies tremor using a two-axis approach based on clinical features and etiology, which are then subclassified into multiple subcategories. Within the category of clinical feature axis and its subcategory of tremor characteristics, tremor is subclassified based on activation characteristics into rest tremor (RT) and action tremor (AT) [2]. The RT has been defined as a tremor in a body part that is not voluntarily activated, assessed when the patient is attempting to relax and is given adequate opportunity to relax the affected body part. Whereas the MDS classification of RT is a singular definition, AT is further sub-divided into postural tremor, kinetic tremor and isometric tremor. AT has been defined as a tremor occurring in a body part while voluntarily maintaining a position against gravity (postural tremor), during any voluntary movement (kinetic tremor), or during muscle contraction against a rigid stationary object (isometric tremor) [2].

Although RT is one of the cardinal features of Parkinson’s Disease (PD), PD patients are also observed to have postural tremor, kinetic tremor or both [3, 4]. In addition to being one of the most visual symptoms of PD, tremor is also ranked as one of the most troubling symptoms by PD patients[5]. Moreover, tremor impairs several physical and psychological quality of life domains in PD patients, similar to essential tremor patients as assessed with Quality of Life in Essential Tremor (QUEST) questionnaire[6]. Importantly, presence of tremor is also used to classify PD into tremor-dominant (TD), indeterminate (IND) and postural instability and gait difficulty (PIGD) subtypes[7], which have been linked with different rates of disease progression [8]. Specifically, a total of 8 and 11 tremor items from Unified Parkinson Disease Rating Scale (UPDRS) or MDS-UPDRS scales are used to calculate tremor score, which is then used in conjunction with the postural instability/gait difficulty (PIGD) score, for computing these subtypes[7].

Despite such significance of tremor in PD, descriptions of prevalence of basic tremor types (RT, postural tremor, kinetic tremor) and relationship of RT with AT in PD are currently limited [3, 9-12], as summarized in table 1. Here, we aimed to describe the prevalence of basic tremor types, and the relationship of RT with AT in three large cohorts of PD patients, including the Parkinson Progression Marker Initiative (PPMI)[13], The Fox Investigation for New Discovery of Biomarkers (BioFIND)[14], and the Parkinson’s Disease Biomarkers Program (PDBP) [15].

**Table 1:** Summary of previous studies reporting tremor prevalence Not reported (NR), Unified Parkinson Disease Rating Scale (UPDRS), Washington Heights-Inwood Genetic Study of ET (WHIGET), Parkinson’s Disease Society Brain Bank (PDSB). The study by Pasquini et. al. used the Parkinson Progression Marker Initiative (PPMI) data and also reported on prevalence of difference tremor types at 2-years follow-up as following: any tremor 83.9%, rest tremor 67.9%, postural tremor 49.5%, and kinetic tremor 46.8%.

| **Variable** | **Rajput**<br>**et al.**<br>**1991** | **Hughes**<br>**et al.**<br>**1993** | **Louis**<br>**et al.**<br>**2001** | **Gigante**<br>**et. al.**<br>**2014** | **Pasquini**<br>**et al.**<br>**2018** |
| --- | --- | --- | --- | --- | --- |
| Study design | Longitudinal,<br>neuropathologic | Longitudinal,<br>neuropathologica | Cross-sectional<br>clinical | Cross-sectional<br>clinical | Longitudina<br>clinical |
| Disease duration<br>(years) | NR | 13.1 ± 6.3 | 7.9 ± 5.1 | 5.2 ± 3.8 | 2.56 ± 0.56 |
| Tremor<br>assessment<br>method | Visual inspectio | PDSBB form | WHIGET<br>Tremor<br>Rating<br>Scale | UPDRS | MDS-UPDR |
| Sample size | 30 | 100 | 197 | 237 | 378 |
| Any tremor<br>prevalence | NR | 69% | NR | NR | 87.8% |
| Rest tremor<br>prevalence | 100% | NR | NR | 83.12% | 69.6% |
| Action tremor<br>prevalence | NR | NR | 93.4% | 46% | NR |
| Postural tremor<br>prevalence | NR | NR | NR | NR | 52.1% |
| Kinetic tremor | NR | NR | NR | NR | 51.6% |
| prevalence |  |  |  |  |  |

## Methods

The PPMI, BioFIND and PDBP are multi-center, observational studies, designed to accelerate research in PD, especially biomarkers discovery, through collection of clinical, biospecimen and other relevant data, such as imaging, by making the data available to researchers in an open-access manner through their respective portals. The PPMI is an international, longitudinal study of early stage PD patients (n= 423), recruited within 6 months from initial diagnosis and not on dopaminergic treatment, and healthy controls (n = 196). The BioFIND is a cross-sectional study of moderate to advanced stage PD patients (n = 118) on dopaminergic treatment and healthy controls (n = 88). The PDBP is a longitudinal study of primarily early to advanced stages PD patients (n= = 882), with most patients on dopaminergic treatment and healthy controls (n= = 549), in addition to patients with other related clinical diagnosis (n= = 168, e.g., atypical parkinsonian disorder, essential tremor). A summary of relevant demographics and clinical features of these three cohorts is presented in table 2, while more details have been described in the respective cohorts elsewhere [13-15].

**Table 2:** Summary of relevant demographics and clinical features of the PPMI, BioFIND and PDBP cohorts. Parkinson Progression Marker Initiative (PPMI), The Fox Investigation for New Discovery of Biomarkers (BioFIND) and Parkinson’s Disease Biomarkers Program (PDBP). * this value was taken from original description of the PDBP cohort for 449 Parkinson’s disease subjects.

| Variable | PPMI (n=423) | BioFIND (n=118) | PDBP (n=874) |
| --- | --- | --- | --- |
| Gender (Male / Female) | 146 / 277 | 62.7% | 563 / 319 |
| Age (mean ± standard deviation) | 61.6 ± 9.7 | 68.0 ± 6.5 | 64.3 ± 9.1 |
| Disease duration | 5.85 ± 7.5 | 8.5 ± 3.2 | 7.2 ± 7.5* |
| % on Dopamine Treatment | 0 | 100% | 90.8% |

We chose to compare the PPMI, BioFIND and PDPB cohorts of PD patients for this study for a variety of reasons: large number of PD patients in each cohort; subjects recruited in each cohort by movement disorders specialists; data were available on open-access basis; and, a wide range of severity of disease, ranging from drug-naïve PD patients in the PPMI cohort, moderately advanced PD patients on dopaminergic treatment (with MDS-UPDRS captured in on-state on baseline visit and off-state in follow-up visit) in the BioFIND cohort, and all stages of PD severity in cohort with majority being on dopaminergic treatment (with MDS-UPDRS irrespective of off-state or on-state) in the PDBP cohort.

Baseline visit data of the PD patients (n = 423 for PPMI, n = 118 for BioFIND, n = 873 for PDBP) were accessed as of January 22, 2020 from the PPMI (https://www.ppmi-info.org) and Accelerating Medicine Partnership in Parkinson’s Disease (AMP PD) databases (https://amp-pd.org/). The For the BioFIND cohort, the MDS-UPDRS part III data from baseline visit (on-state) were used by default for all calculations, while the follow-up visit data (off-state; 14 days after the baseline visit) were used for comparison separately.

The three basic tremor types, specifically, RT, postural tremor, and kinetic tremor were captured by value of >= 1 on items 3.17 (*rest tremor amplitude*), 3.15 (*postural tremor of the hands*), and 3.16 (*kinetic tremor of hands*), respectively, from part III (motor) of the MDS-UPDRS in all three cohorts. For the purpose of this study, we defined AT to include both postural and kinetic, rather than postural or kinetic tremor, as former approach yields higher specificity and avoids false positives, given that re-emergent rest tremor can be frequently mistaken as postural tremor[16]. We also defined RT severity (range 0 – 80) by adding five sub-items of RT (3.17) and then multiplying the sum with item 3.18 (*constancy of rest tremor*) (3.18) from part III (motor) of the MDS-UPDRS. We also assessed the perception of tremor by patient using item 2.10 (*Over the past week, have you usually had shaking or tremor?*) from part II of the MDS-UPDRS.

We first analyzed the average prevalence of RT (irrespective of postural or kinetic tremor), pure RT (with neither postural tremor or kinetic tremor), AT (irrespective of RT), pure AT (with no RT), postural tremor (irrespective of RT or kinetic tremor), pure postural tremor (with neither RT or kinetic tremor), kinetic tremor (irrespective of RT or postural tremor), pure kinetic tremor (with neither RT or postural tremor), no tremor (neither RT, postural tremor or kinetic tremor), any tremor (any of three basic tremor types) and all tremor (all three basic tremor types), and distribution of perception of tremor across baseline data of the three cohorts and in each individual cohort. We then tested the hypothesis that prevalence RT and AT are not independent of each other, such that the severity of RT would be higher in PD patients with AT versus without AT, in individual cohorts. Finally, we also compared prevalence of RT, AT, postural tremor, kinetic tremor, any tremor and all tremor in off-state and on-state in the BioFIND cohort. Statistical testing was performed using SPSS version 26 with appropriate statistical tests at a significance level (p < 0.05).

## Results

The average prevalence of RT and pure RT was 58.2% and 14.5 %, respectively. In contrast, the average prevalence of AT and pure AT were 36.6% and 9.6%, respectively. The average prevalence of postural tremor and pure postural tremor was 49.7% and 4.0 %, respectively. The average prevalence of kinetic tremor and pure kinetic tremor was 52.3% and 8.1 %, respectively. The average prevalence of patients with no tremor, any tremor and all tremor were 19.9%, 79.9%, and 26.9%, respectively. Figure 1 provides a summary of these results using a diagram. Table 3 summarizes these results in each individual cohort.

**Table 3:** Distribution of prevalence of different types and combinations of tremor in each of three cohorts. Parkinson Progression Marker Initiative (PPMI), Fox Investigation for New Discovery of Biomarkers (BioFIND), Parkinson’s Disease Biomarkers Program (PDBP).

| **Tremor Type** | **PPMI (n=423)** | **BioFIND (n=118)** | **PDBP (n=874)** |
| --- | --- | --- | --- |
| Rest tremor | 290 (68.6%) | 75 (63.6%) | 459 (52.0%) |
| Pure rest tremor | 87 (20.6%) | 15 (12.7%) | 104 (11.8%) |
| Action tremor | 156 (36.9%) | 46 (39.0%) | 316 (35.8%) |
| Pure action tremor | 40 (9.5%) | 10 (8.5%) | 87 (9.9%) |
| Postural tremor | 223 (52.7) | 69 (58.5%) | 412 (46.7%) |
| Pure Postural tremor | 18 (4.3%) | 8 (6.8%) | 31 (3.5%) |
| Kinetic tremor | 217 (51.3%) | 61 (51.7%) | 463 (52.5%) |
| Pure kinetic tremor | 23 (5.4%) | 6 (5.1%) | 86 (9.8%) |
| No tremor | 52 (12.3%) | 19 (16.1%) | 211 (23.9%) |
| Any tremor | 371 (87.7%) | 99 (83.9%) | 663 (75.2%) |
| All tremor | 116 (27.4%) | 36 (30.5%) | 229 (26.0%) |

**Figure 1:**
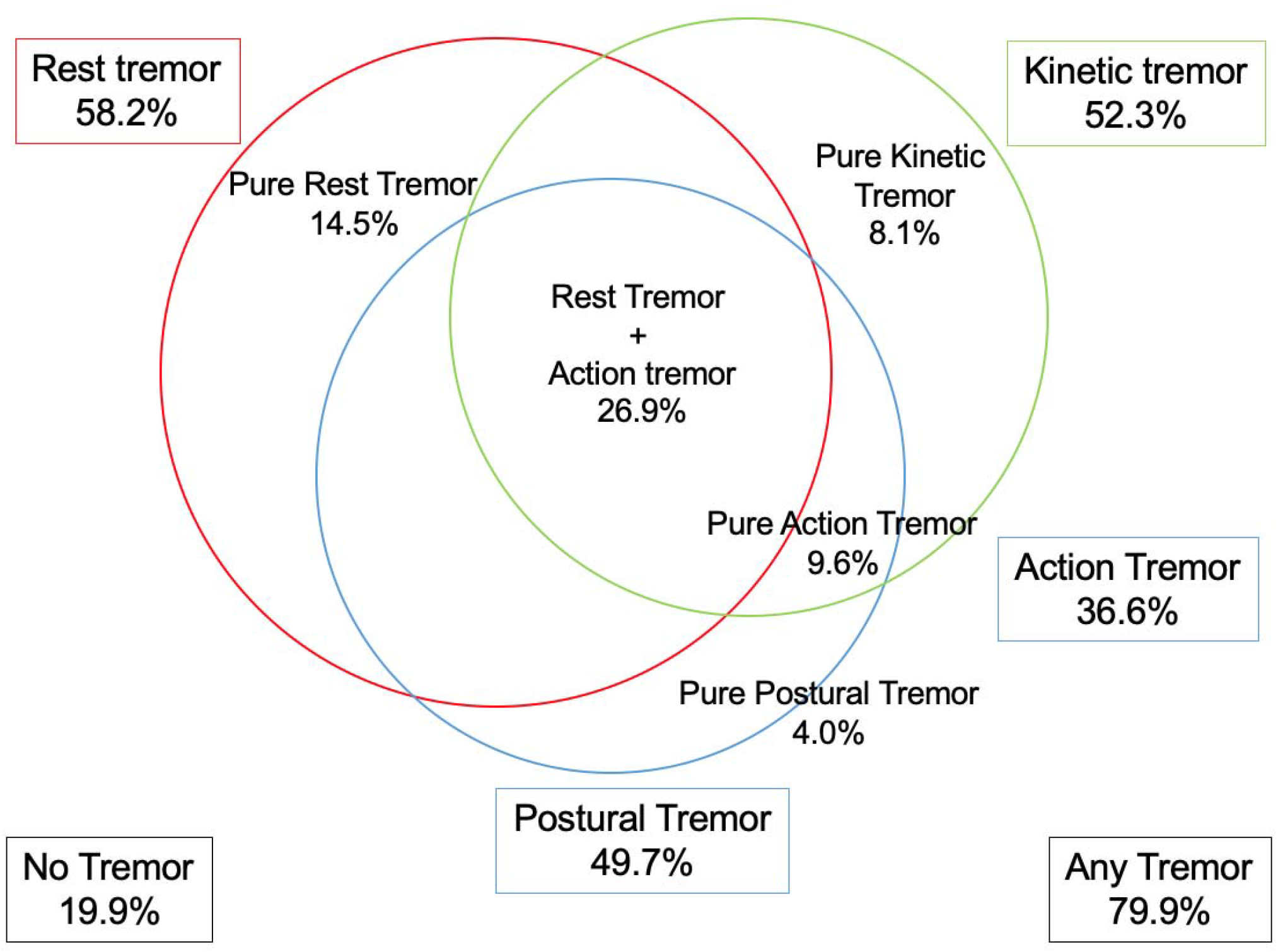
Average prevalence of different tremor types and their combinations across baseline data of the PPMI, BioFIND and PDBP cohorts. Parkinson Progression Marker Initiative (PPMI), The Fox Investigation for New Discovery of Biomarkers (BioFIND) and Parkinson’s Disease Biomarkers Program (PDBP). Size of the circles are not indicative of the prevalence magnitude.

As for average perception of tremor by the patient over past week, 19.5% of the patients reported having no tremor, while 52.8%, 21.4%, 5.3% and 1.0%, reported having slight, mild, moderate and severe tremor, respectively. Table 4 summarizes these results in each individual cohort. Conversely, on average 79.5% patients reported having any shaking or tremor (slight, mild, moderate or severe) over past week.

**Table 4:** Distribution of perception of tremor in each of three cohorts. Movement Disorders Society Unified Parkinson’s Disease Rating Scale (MDS-UPDRS).

| Variable | PPMI (n=422) | BioFIND (n = 118) | PDBP (n=826) |
| --- | --- | --- | --- |
| No tremor | 59 (14.0%) | 24 (20.3%) | 183 (22.1%) |
| Subtle tremor | 282 (66.8%) | 59 (50.0%) | 380 (46.0%) |
| Mild tremor | 73 (17.3%) | 30 (25.4) | 189 (22.9%) |
| Moderate tremor | 7 (1.7%) | 4 (3.4%) | 62 (7.5%) |
| Severe | 1 (0.2%) | 1 (0.8%) | 12 (1.5%) |

We also found that patients with RT versus without RT consistently had a significantly higher (Chi-square test) proportion of AT in the PPMI (40.0% versus 30.1%, p = 0.049), in the BioFIND (48.0% versus 40.0%, p = 0.008) and PDBP (49.9% versus 21.0%, p = 0.000) cohorts, respectively. Based on these observations, we postulated that PD patients might develop AT after RT becomes severe, or conversely, PD patients with AT would have higher severity of RT compared to patients without AT. In support of this hypothesis, we found that the severity of RT was significantly higher (Student’s t-test) in patients with AT than patients without AT in the PPMI (5.7 ± 5.4 versus 3.9 ± 3.3, p = 0.000), BioFIND (6.4 ± 6.3versus 3.8 ± 4.4, p = 0.000) and PDBP (6.4 ± 6.6 versus 3.7 ± 4.4, p = 0.000) cohorts, respectively.

In the BioFIND cohort (in which patients were examined in on-state on baseline visit and off-state in second visit), we found that the prevalence of RT, AT, postural tremor, kinetic tremor, any tremor and all tremor were significantly (p < 0.05, Chi-square test) higher in off-state, compared to on-state (figure 2). The RT severity was also higher in off-state (5.87 ± 8.94) compared to on-state (4.04 ± 6.76), however, this difference did not reach statistical significance (p=0.051).

**Figure 2:**
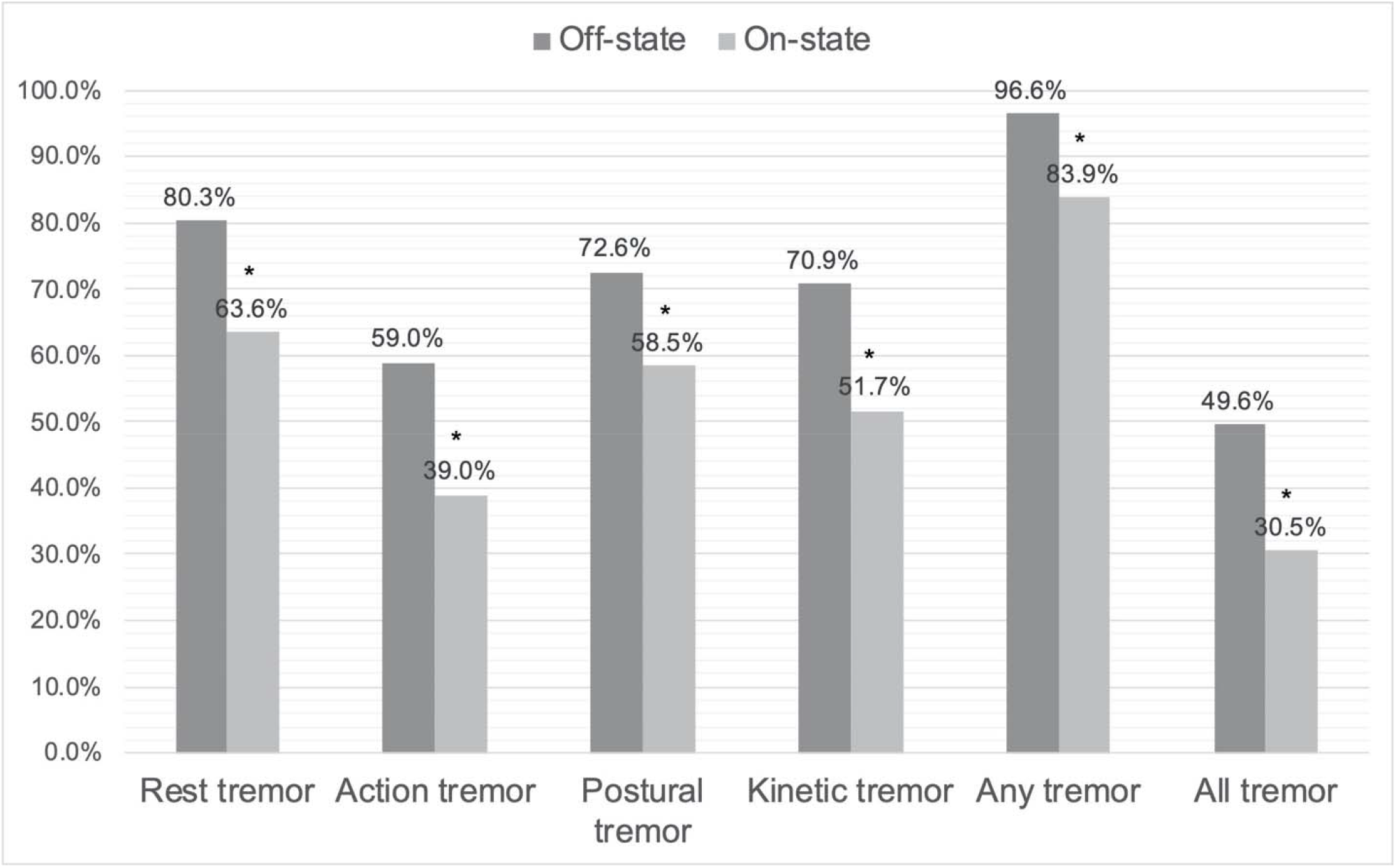
Changes in tremor prevalence from off-state to on-state in the BioFIND cohort. Fox Investigation for New Discovery of Biomarkers (BioFIND). On-state data were obtained from the baseline visit; off-state data were obtained by the follow-up visit (14 days after the baseline). *p<0.05, Chi-Square test.

## Discussion

Our study results indicate that across the PPMI, BioFIND and PDBP cohorts, although RT has the highest prevalence (58.2%) in PD, AT can also be present in a sizable number (36.2%) of PD patients, which is in line with similar literature[11, 12, 17]. In comparison, a large majority (79.5%) of the patients self-report having tremor. Our study results also indicate all tremor types are reduced by dopaminergic treatment, which is consistent with the traditionally held view on the influence of levodopa and dopamine agonist on tremor in PD[18, 19].

We also found that the PD patients with RT had significantly higher prevalence of AT in all three cohorts. Moreover, we found that the patients with AT, compared to patients without AT, had more severe RT in all three cohorts, similar to findings reported in previous studies[11, 12]. These findings together demonstrate that AT could be a part of broader tremor syndrome of PD. Furthermore, these findings are supported by functional neuroimaging and electrophysiology studies demonstrating cross-interactions between basal ganglia and cerebellothalamic circuitry in mediating tremor in PD [20, 21]. The presence of re-emergent RT or a comorbid alternative tremor disorder (such as essential tremor) could, at least in part, explain these findings. It is worth noting that MDS-UPDRS scale does not distinguish between postural tremor and re-emergent rest tremor mimicking postural tremor, and such distinction can only definitely be made with electrophysiological analysis [16].

One of the limitations of our study was that the data used for defining prevalence of different types of tremor were collected by multiple investigators in the PPMI, BioFIND and PDBP cohorts. However, these data are considered valid as the clinical examination for recording presence of tremor were done as part of the standardized MDS-UPDRS by trained movement disorders specialists in all three of these studies conducted at internationally recognized movement disorders centers. Another limitation was that we could not include isometric tremor in defining AT, which was unavoidable since the MDS-UPDRS scale does not capture isometric tremor. It is also worth noting that MDS-UPDRS does not capture postural and kinetic tremor in lower limbs or head, unlike RT, which is captured in upper limbs, lower limbs and lip / jaw.

The presence of RT is not mandatory for a diagnosis of PD, which is supported by our data demonstrating absence of any tremor and presence of pure AT in nearly one-fifth and one-tenth of all patients across the PPMI, BioFIND and PPMI cohorts. These observations have implications in clinical practice and research enrollment, where the presence of patients either without tremor or with multiple tremor subtypes potentially lead to diagnostic delay, ambiguity and uncertainty. For example, different patterns of basic tremor types have been shown to improve differential diagnosis of PD from essential tremor[22]. All together, these results provide further evidence that PD tremor is highly heterogenous and its correct phenotypic classification in PD will be essential in optimizing diagnostic, therapeutic and prognostic approaches.

## Data Availability

Data used in this study are available at the respective portal.

https://www.ppmi-info.org/

https://biofind.loni.usc.edu/

https://amp-pd.org/

## Acknowledgement

PPMI – a public-private partnership – is funded by the Michael J. Fox Foundation for Parkinson’s Research and funding partners, including AbbVie, Allergan, Avid, Biogen, BioLegend, Bristol-Myers Squibb, Celgene, Denali, GE Healthcare, Genetech, GSK, Lilly, Lundbeck, Merk, MSD, Pfizer, Piramidal, Prevail, Roche, Sanofi Genzyme, Servier, Takeda, Teva, UCB, Verily, Voyager, Golub Capital.

Data used in the preparation of this article were also obtained from the AMP PD Knowledge Platform. For up-to-date information on the study, https://www.amp-pd.org.” AMP PD – a public-private partnership – is managed by the FNIH and funded by Celgene, GSK, the Michael J. Fox Foundation for Parkinson’s Research, the National Institute of Neurological Disorders and Stroke, Pfizer, Sanofi, and Verily.”

